# Neuromodulation through Brain Stimulation-assisted Cognitive Training in Patients with Post-Chemotherapy Cognitive Impairment (Neuromod-PCCI): Study Protocol of a Randomized Controlled Trial

**DOI:** 10.1101/2022.04.04.22273380

**Authors:** Merle Rocke, Elena Knochenhauer, Friederike Thams, Daria Antonenko, Anna Elisabeth Fromm, Nora Jansen, Ulrike Grittner, Sein Schmidt, Eva-Lotta Brakemeier, Agnes Flöel

**Author notes:** Corresponding author: Agnes Flöel, MD, University Medicine Greifswald, Department of Neurology Ferdinand-Sauerbruch-Straße, D-17475 Greifswald. contributed equally (Merle Rocke, Elena Knochenhauer).

## Abstract

**Introduction:** Breast cancer is the most common form of cancer in women. A considerable number of women with breast cancer who have been treated with chemotherapy, subsequently develop neurological symptoms such as concentration and memory difficulties (also known as ‘chemobrain’). Currently, there are no validated therapeutic approaches available to treat these symptoms. Cognitive training holds the potential to counteract cognitive impairment. Combining cognitive training with concurrent transcranial direct current stimulation (tDCS) could enhance and maintain the effects of this training, potentially providing a new approach to treat post-chemotherapy cognitive impairment (PCCI). With this study, we aim to investigate the effects of multi-session tDCS over the left dorsolateral prefrontal cortex in combination with cognitive training on cognition and quality of life in women with PCCI.

**Methods and analysis:** The Neuromod-PCCI trial is a monocentric, randomized, double-blind, placebo-controlled study. Fifty-two women with PCCI after breast cancer therapy will receive a 3-week tDCS-assisted cognitive training with anodal tDCS over the left dorsolateral prefrontal cortex (target intervention), compared to cognitive training plus sham tDCS (control intervention). Cognitive training will consist of a letter updating task. Primary outcome will be the performance in an untrained task (n-back task) after training. In addition, feasibility, safety and tolerability, as well as quality of life and performance in additional untrained tasks will be investigated. A follow-up visit will be performed one month after intervention to assess possible long-term effects. In an exploratory approach, structural and functional magnetic resonance imaging (MRI) will be acquired before the intervention and at post-intervention to identify possible neural predictors for successful intervention.

**Ethics and dissemination:** Ethical approval was granted by the ethics committee of the University Medicine Greifswald (BB236/20). Results will be available through publications in peer-reviewed journals and presentations at national and international conferences.

**Trial registration:** This trial was prospectively registered at ClinicalTrials.gov (Identifier: NCT04817566, registered March 26, 2021).

**Strength and limitations of this study:**

– This is the first randomized controlled trial to investigate the feasibility and effects of combined cognitive training and tDCS on cognitive outcomes and quality of life in patients with post-chemotherapy cognitive impairment
– Results will help the development of treatment options for breast cancer patients with post-chemotherapy cognitive impairment
– Results may not be generalizable to male cancer patients
– Monocentric trial design may increase risk of bias

## Introduction

With the development of new treatment options, the long-term survival rates of breast cancer patients is increasing ^1^. However, cancer patients may develop cognitive impairments following chemotherapy ^2 3^. Post-chemotherapy cognitive impairment (PCCI), also referred to as “chemobrain”, can persist for many years after chemotherapy and occurs in various cognitive domains, such as executive functions, memory, processing speed, and attention ^3 4^. Beyond that, cognitive impairment may cause psychological distress and affect quality of life ^5-8^.

The pathophysiology of PCCI remains poorly understood. Multiple mechanisms underlying patterns of cognitive impairment in cancer patients have been proposed, including neuroinflammation, direct neurotoxicity, as well as alterations in structure, function and metabolic profile of the brain ^9^ as well as changes in neurotransmitters, such as reduced concentrations of norepinephrine, dopamine and serotonin, among others ^10^.

Thus far, no evidence-based treatment options are available. First studies have found preliminary evidence that cognitive training may improve cognitive abilities of patients with PCCI ^11-13^. Von Ah et al. observed an improvement in trained functions and a transfer effect to untrained memory tasks as well as to self-rated subjective cognitive function, and reduced psychological stress in the trained group compared to a waiting list control group after training of memory and speed of processing ^12^. Similarly, in the study of Kesler et al. participants trained cognitive flexibility, working memory, processing speed and verbal fluency, and subsequently showed enhanced performance in cognitive functions meditated by the prefrontal cortex as well as minor improvements in declarative memory scores^13^. Despite these promising preliminary findings, training programs have been time-intensive, involving several weeks of task practice, and transfer effects have not consistently been demonstrated^14^.

Non-invasive brain stimulation is an approach to boost training gains, which allows for shorter training periods and for induction of transfer to untrained functions. Transcranial direct current stimulation (tDCS) is an inexpensive, painless method that is well-tolerated, safe and easy to apply ^13^. This promising approach is already widely used for interventional and therapeutic purposes (see ^15 for review^). For administration of tDCS, a weak electric current is applied via two or more electrodes on the participant’s scalp. The supposed mechanism of action is a directional modulation of the cortical excitability in the brain area under the electrodes as well as in functionally and structurally connected brain regions ^16^. The application of direct current causes a shift in the resting membrane potential of the cortical neurons which in turn leads to a decrease of the threshold for triggering action potentials. Accordingly, the corresponding area of the brain shows a higher level of excitability. Furthermore, the excitatory effect of anodal stimulation leads to an increase in synaptic transmission, which can last for hours to days. This effect corresponds to long-term potentiation-like mechanism and can induce neural plasticity ^17^. Promising results of the technique with regard to improving cognitive performance have been demonstrated in healthy aging, but also in patients with dementia or mild cognitive impairment (MCI) ^18 19^. To our knowledge, no study to date has examined the effects of tDCS-assisted cognitive training in patients with PCCI in order to establish its potential to beneficially impact cognitive functions and patient-reported outcome measures such as health-related quality of life (HRQoL). In the Neuromod-PCCI study, we plan to assess in a double-blind randomized controlled phase II clinical trial if such a combined multi-session cognitive training plus tDCS intervention yields substantial long-term benefits and transfer effects in patients with PCCI. All patients will complete cognitive training of a letter updating task over nine training sessions with concurrent tDCS over the left dorsolateral prefrontal cortex (dlPFC). Half of the sample will receive anodal tDCS while performing the cognitive training, whereas the other half will undergo sham stimulation during training. The intervention will span three weeks, with three training sessions per week. We will assess behavioral outcome measures, such as direct training effects, feasibility, safety and tolerability, transfer to untrained domains, and long-term effects on cognition and quality of life at multiple time points. We will further elucidate neural characteristics associated with interventional success before the intervention and assess neural correlates at post-intervention. This protocol, describing the design and methods of the Neuromod-PCCI study, was prepared in accordance with the SPIRIT guidelines ^20 21^

## Methods: Participants, Intervention, and Outcomes

### Design and Setting

This is a monocentric, randomized, double-blind, placebo-controlled study. The intervention is comprised out of nine sessions of cognitive training over three weeks, accompanied by anodal tDCS over the left dlPFC compared to sham tDCS. Subjects will participate in 13 sessions altogether with pre- and post-intervention assessments. One follow-up session is planned one month after the intervention to assess possible long-term effects. All sessions will take place at the University Medicine Greifswald, Greifswald, Germany. Magnetic resonance imaging (MRI) will be performed before the intervention and at post-intervention (“add-on” study, i.e. optional for the participants). A flowchart of the study is shown in **Figure 1**.

**Figure 1:**
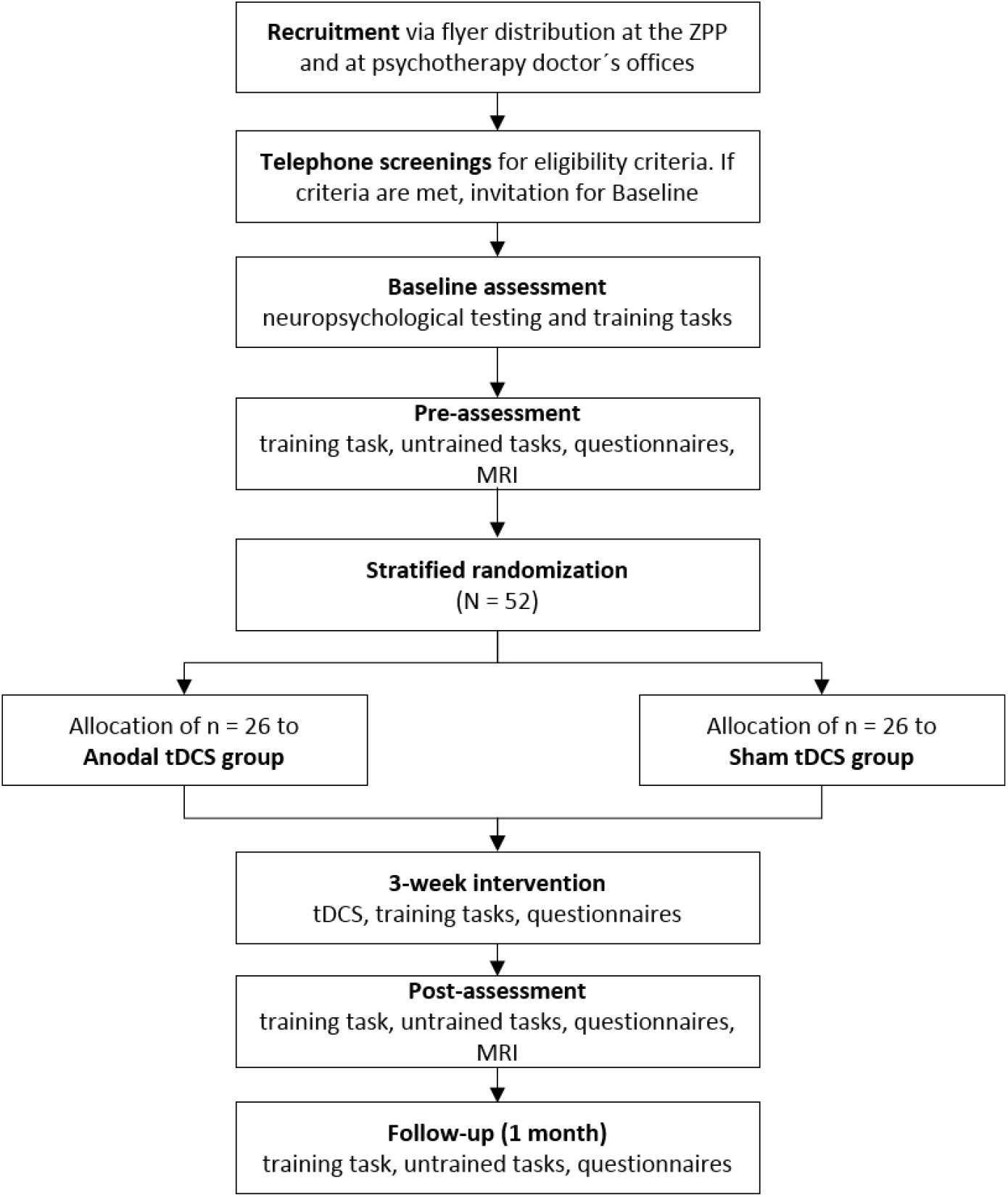
Neuromod-PCCI study flowchart. MRI, magnetic resonance imaging. tDCS, transcranial direct current stimulation. ZPP, Zentrum für Psychologische Psychotherapie (university psychotherapy outpatient clinic).

### Eligibility Criteria

Participants included in the study must meet all of the following inclusion criteria:

- Chemotherapy to treat breast cancer (over 6 months since end of treatment)
- Self-reported concerns regarding cognitive function
- Age: 18-65 years
- Right-handedness

In case one or more of the following criteria are present at randomization, potential participants will be excluded:

- History of dementia before treatment of cancer
- Other neurodegenerative neurological disorders, epilepsy or a history of seizures
- Severe and untreated medical conditions that preclude participation in the training, as determined by responsible physician
- History of moderate to severe substance use disorder according to DSM-5 ^22^
- Moderate to severe acute mental disorders according to DSM-5 ^22^
- Contraindication to tDCS ^23^

Note that contraindication to MRI will not be treated as exclusion criteria as participants will still be included in the study sample, but no MRI scans will be performed in these individuals. If all eligibility criteria are met and participants provide written informed consent, they will be included in the study sample.

### Intervention

At each of the training sessions participants will receive either anodal or sham tDCS while completing a cognitive training task. Before starting the training task, the scalp of the participants will be numbed via an application of a topical anesthetic (EMLA® Cream, 25mg/g Lidocain + 25 mg/g Prilocain). The EMLA cream will be applied on the scalp according to the positioning of the electrodes and will be left to take effect for about 20 min. After the exposure time, the EMLA cream will be removed, the tDCS setup will be mounted and stimulation will be started.

Participants will be presented with a letter updating (LU) task on a tablet computer to train working memory updating. List of letters A to D (with lengths of 5, 7, 9, 11, 13 or 15 letters, six times each; total of 36 lists) will be presented in random order, one letter at a time (presentation duration 2000 ms, ISI 500 ms). After each list participants will be asked to recall the last four letters that were presented.

TDCS will be administered via a battery-operated stimulator (Neuroelectrics Starstim 8, Barcelona, Spain). A current of 2 mA will be applied via five round electrode-gel filled NG PigStim electrodes (NE029, Neuroelectrics, Barcelona, Spain), connected to the stimulator and mounted in a neoprene head cap using the 10-20 EEG grid. The stimulation will be applied continuously for 20 min, with 20 additional sec of ramping at the beginning and at the end of stimulation, respectively. The anodal electrode will be placed over the left dlPFC (F3). The four cathodal electrodes will be arranged in a circle around the anodal electrode, in the positions of AFz, F7, C5 and FCz respectively, to constrain the current flow to the target region. In the sham group, current will be applied for 30 sec to blind the participants regarding their stimulation condition. Ramp times and electrode montage will be the same as in the anodal group. The cognitive training task and the stimulation will be started simultaneously. After every third session, participants will fill out an adverse-events questionnaire ^23^. Participants will be instructed to avoid excessive consumption of alcohol or nicotine on the days of intervention. Moreover, they will be asked to adhere to their regular sleep schedule and to avoid drinking caffeine 90 min prior to the training sessions.

### Outcome Measures

#### Primary Outcomes

Primary outcome measure will be working memory performance at post-assessment, operationalized by the percentage of correct responses in the n-back task (untrained task).

#### Secondary Outcomes

Secondary outcome will be working memory performance in the untrained task (nback, ^24^) at follow-up assessment, operationalized by the percentage of correct responses. Additionally, performance in primary memory training task (LU task, ^25^) and visuospatial memory performance (virtual reality (VR) task, ^26^) will be assessed at post- and follow-up-assessments. Performance in the training task will be operationalized by number of correctly recalled lists in the LU task and visuospatial performance will be operationalized by number of correctly recalled items in the VR task.

Further secondary outcome measures include Quality of Life, operationalized via the PROMIS Preference score (PROPr score) which serves as a summary scoring system of health-related quality of life including domains such as depression, fatigue, pain interference and ability to participate in social roles and activities^27^. The domain of cognitive function will be scored separately because it is the most important subscale for this study. Scoring will be carried out according to the scoring manuals (http://www.healthmeasures.net/). Feasibility will be operationalized through drop-out rates (% participants that did not complete post-assessment), adherence rates of training sessions (number of training sessions attended) and the post-study system usability questionnaire (PSSUQ)^28^, completed by study assessors. Additionally, structural and functional neural correlates (assessed at pre- and post-assessments), as measured by structural and functional MRI, will be assessed.

#### Exploratory Analyses

Exploratory analyses will be conducted for more detailed outcomes of the training task (e.g., outcomes dependent on list length in the LU task). Additionally, education, baseline cognitive ability or neuropsychological status as well as socio-demographic and clinical characteristics will be analyzed to identify potential predictors of training task performance and response to the intervention.

#### Participant Timeline

Individuals will participate in 13 visits, and two optional MRI sessions, taking place at the University Medicine Greifswald. After inclusion at baseline visit (V0), participants will attend the pre-assessment visit (V1) before starting the nine training visits during three consecutive weeks on three days a week (V2-V10). After the training, a post-assessment (V11) will be conducted and four weeks later a follow-up visit (V12) will be administered. MRI will be acquired before pre-assessment (V1) and at post-assessment (V13).

#### Baseline Measures

At baseline, participants will provide written informed consent and participate in a demographic interview. The participants will be asked to provide a record of their cancer specific medical history. If study-relevant information is missing, it will be requested from the responsible physician. Furthermore, a diagnostic interview to screen for mental disorders will be conducted and subsequently the participants will be tested with a comprehensive battery of neuropsychological tests to quantify cognitive function on different domains. The following interviews, questionnaires and tests will be conducted:

- Diagnostic interview for mental disorders (semi-structured diagnostic interview, DIPS)^29^ to screen amongst others for moderate to severe acute mental disorders according to DSM-5 ^22^ including depression, anxiety disorders, post-traumatic stress disorder and sleep disorders
- Childhood Trauma Screener (CTS)^30^ to screen for childhood mal-treatment
- German version of the Auditory verbal learning test (AVLT)^24 31^ to test verbal memory
- Rey-Osterrieth Complex Figure Test (ROCF)^32^ to test visual-spatial memory
- Digit Span ^24^ and Trail Making Test A/B (TMT A/B)^33^ to test short term/working memory
- Stroop test ^24 34^ and Semantic word fluency test ^24^ to test executive functions

Afterwards, participants will perform the training task as described above, with the exception that at baseline the task will consist of one practice trail with 4 lists and one actual training trial with 25 lists (compared to 36 lists in the training). The baseline assessment will have an approximated duration of 3,5h.

An actigraph (GT3X, ActiGraph, Pensacola, FL, USA) will be handed out to the participants at baseline to record sleep and activity data. Participants will be instructed to wear the device in the week leading up to pre-assessment and in the week following the post-assessment. Additionally, at baseline, participants will be asked to complete two sleep questionnaires: the Epworth Sleepiness Scale to measure daytime sleepiness ^35^ and the Morningness-Eveningness-Questionnaire ^36^ to estimate the participant’s circadian rhythm.

#### Pre-, Post-, and Follow-Up-Assessments

All three sessions will follow the same procedure. First, self-reported well-being, quality and duration of sleep as well as potential stressors up to two hours prior to the visit will be assessed by the investigator via a questionnaire. The participants will further complete an array of QoL-related questionnaires: Patient Reported Outcomes Measure System (PROMIS®) which evaluates physical, mental and social health ^27^ and the EORTC QLQC30 which assesses the quality of life in cancer patients, supplemented with the module EORTC BR23 with the focus on breast cancer ^37^. To further complete the data assessment regarding sleep, the Pittsburgh Quality Sleep Index ^38^ will be administered to assess sleep quality over a longer interval. Then, participants will complete the training task and untrained tasks. The follow-up assessments will provide the possibility for assessing the maintenance of training and transfer effects.

#### Sample Size

Power calculation is based on recent studies using multi-session application of cognitive training compared to a control training on immediate performance in the trained task (primary outcome) ^39-41^. Based on these data, we estimated an effect size of 0.8 (Cohen’s d). To demonstrate an effect in the primary outcome between cognitive training groups and control (% correct in the n-back task for the anodal vs. sham stimulation group) with an independent t-test using a two-sided significance level of α = 0.05 and a power of at least 80 %, 52 participants (26 for target intervention group, 26 for control intervention group) need to be included. This conservative approach using a t-test was chosen for sample size estimation, even though we intend to analyze the primary outcome conducting an analysis of covariance (ANCOVA) model ^42^. This monocentric clinical trial will serve to calculate sample size for a subsequent multicenter clinical trial. Sample size estimation was conducted using R software (http://www.R-project.org) and the pwr package (https://cran.r-project.org/package=pwr).

#### Recruitment

The participants will be recruited by the distribution of flyers at the university psychotherapy outpatient clinic (Zentrum für Psychologische Psychotherapie) and at local psychotherapeutic doctor’s offices in Greifswald and surroundings.

Furthermore, we will search for participants via articles and announcements in the local newspaper. Telephone screenings assessing inclusion and exclusion criteria will be conducted with all potential participants and study information will be provided. Eligible participants will be invited for baseline assessment.

## Methods: Assignment of Interventions

Allocation of the participants to the experimental groups will be performed by a researcher not involved in the study. Participants will be randomly allocated in a 1:1 ratio to the two groups (anodal vs. sham tDCS). Baseline performance in the n-back task will be used as strata (two performance strata; ≤ 87 % correct and > 87 % correct in the n-back task). Randomization blocks with varying block sizes will be generated for each of the two stimulation groups, using R software (https://www.R-project.org) and the blockrand package (http://CRAN.R-project.org/package=blockrand). Participants will then be allocated to the anodal or sham tDCS group based on the generated randomization sequences within each block and stratum.

### Blinding

As this is a double-blind trial, participants and investigators will be blinded regarding the stimulation condition. In the sham group the current will be applied for 30s to blind the participants to the intervention. In previous studies, sham tDCS has been shown as a safe and valid method of blinding participants ^43-46^. At the end of the nine-session intervention, participants will be asked to state if they believe they received anodal or sham stimulation. As for the investigator blinding, a staff member who is not involved in this project will perform the randomization according to the procedure described above and provide unidentifiable labels for the two stimulation protocols (anodal, sham). The investigator involved in this study will select the protocol that corresponds to the participants ID number and will be able to apply the stimulation accordingly without knowledge of the respective stimulation condition.

## Methods: Data Collection, Management, and Analysis

### Data Collection Methods

Neuropsychological, behavioral and sleep data will be collected from each participant. Study investigators will be thoroughly trained in administering the assessments. Additionally, participants will receive an in-depth training on the handling of the Actigraph so that they can adequately and safely administer the sleep recording on their own. The questionnaires that are handed out to the participants will be explained by an investigator and will be accompanied by additional written instruction on how and when to complete the questionnaire. Time points and methods of data collection are shown in **Table 1**. MRI data will be collected for individuals participating in the “add-on” MRI study (optional) unless there are contraindications to MRI scanning.

**Table 1.**
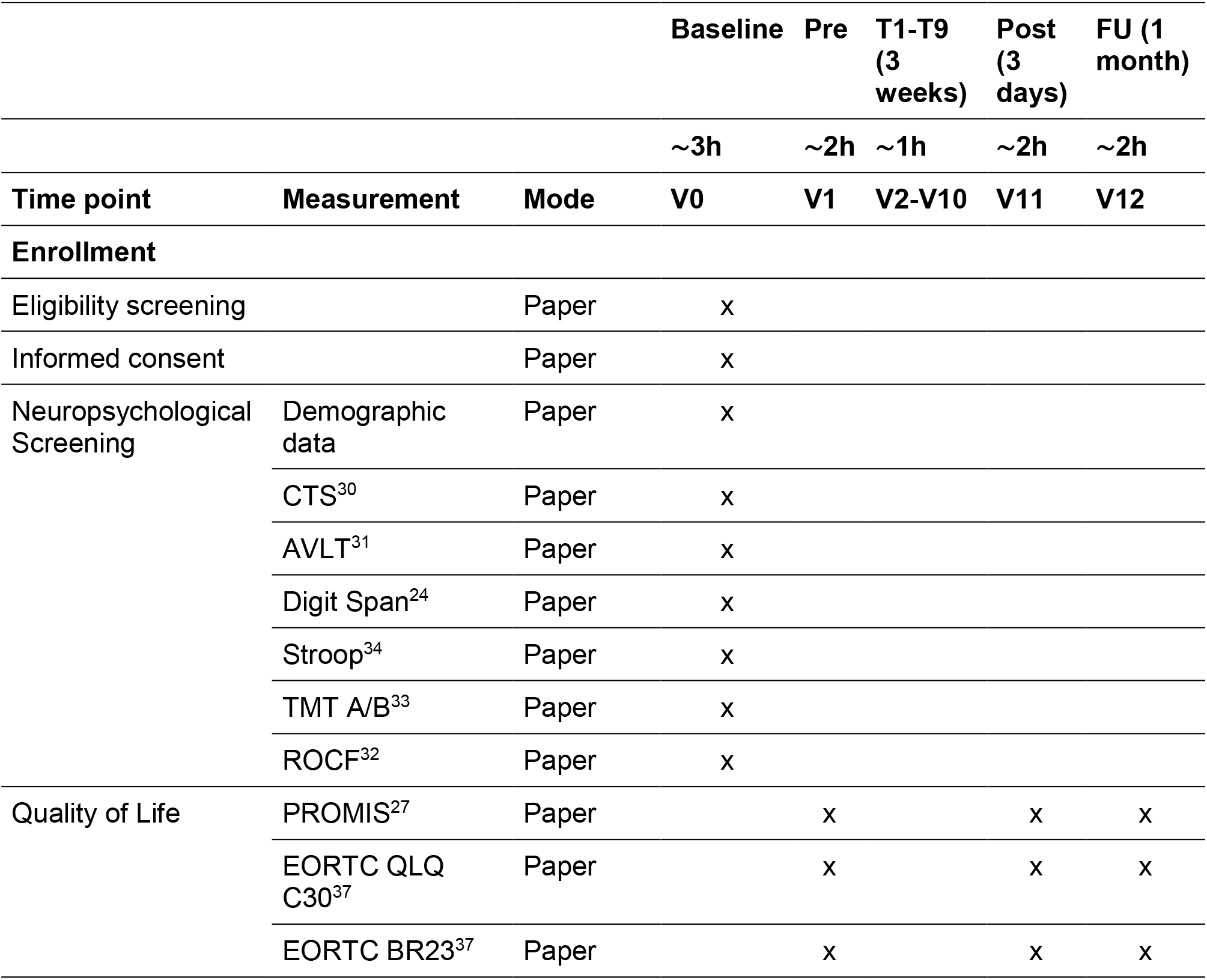
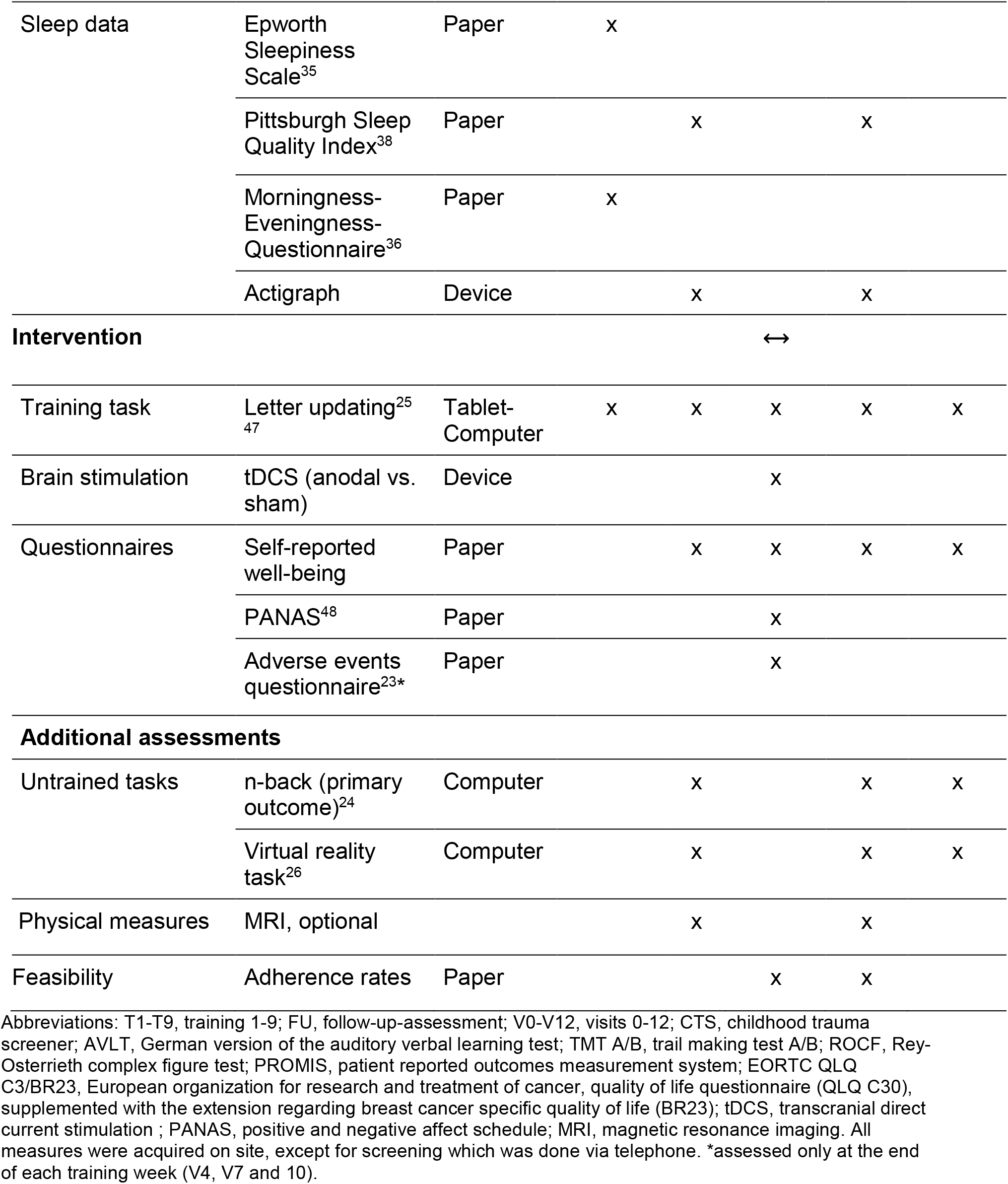
Neuromod-PCCI outcome measures.

### Neuropsychological Assessment

Neuropsychological tests at baseline visit (V0) comprise paper-pencil as well as computer-based assessments. Performance in several cognitive domains will be tested with the auditory verbal learning test^24 31^, the Digit Span^24^, the Stroop test^24 34^, the Trail Making Test A/B ^33^, and the Rey-Osterrieth Complex Figure Test ^32^.

Trained and untrained tasks include paper-pencil and computer-based assessments. Detailed description of the training task is provided in the interventions section. At pre-, post- and follow-up sessions (V1, V11 and V12) the untrained tasks will be administered: Participants will perform a numeric n-back task (1 and 2 back) and a VR navigation task ^26^. The n-back (1-and 2-back) task will be administered as an untrained task to assess working memory function (18 trials total, 9 trials 1back and 9 trials 2back with 10 items each, presentation duration 1500 ms, ISI 2500 ms). The participants will have to state for a sequence of numerical digits presented one after the other, if the stimulus that is presented currently is the same stimulus as “n”-steps back. The participants will answer via a keypress, left arrow key for “yes” if the stimuli match and right arrow key for “no” if the stimuli differ. This task requires that different brain functions are coordinated effectively including updating, discrimination and matching of the stimuli and decision making ^24 49^. The VR task ^26^ will be administered as a untrained task. Here, during encoding, participants will be instructed to memorize a route with several targets (e. g., butcher, doctor’s office and grocery store); during subsequent recall, the participants will be asked to navigate the shortest route to given targets.

### Psychological Assessment

A variety of questionnaires will be administered over the course of the study. At baseline visit (V0) the participants will be screened for childhood maltreatment and various mental disorders via the CTS ^30^ and the DIPS ^29^. The CTS is a 5-item questionnaire regarding experiences of neglect and abuse in childhood and adolescence. The DIPS is an established diagnostic interview which helps to thoroughly explore different possible mental diagnoses. It guides the interviewer, covering the different aspects of each disorder, the degree of symptom expression, and subsequently aids in classifying the possible mental disorders according to DSM-5.

At pre-, post- and follow-up assessment the following Quality of Life questionnaires will be administered:

- Patient Reported Outcomes Measure System (PROMIS®, https://promis-germany.de/) to assess physical, mental and social health
- EORTC QLQC30 ^37^, to measure QoL of cancer patients
- EORTC BR23, which is a module of the EORTC QLQ C30 and focuses on the mental and physical health of breast cancer patients

Simultaneously with the collection of sleep data through actigraphy, the participants will be asked to fill in three questionnaires regarding their sleep quality and rhythm. The Morningness-Eveningness-Questionnaire ^36^ and the Epworth Sleepiness Scale ^35^ will be administered at Baseline assessment, the Pittsburg Sleep Index ^38^ will be administered both at pre-and post-assessment, assessing day-time sleepiness, quality of sleep and the sleep rhythm of the participants in the last month/weeks.

### Magnetic resonance imaging

MRI will be acquired at the Baltic Imaging Center (Center for Diagnostic Radiology and Neuroradiology, Universitätsmedizin Greifswald) with a 3-Tesla scanner (Siemens Verio) using a 32-channel head coil, prior to the training intervention and at post-intervention. A T1-weighted 3D sequence, a 3D FLAIR, a diffusion tensor imaging (DTI), single-voxel magnetic resonance spectroscopy (MRS) and a resting-state fMRI sequence will be recorded. Additional T1- and T2-weighted structural images will be acquired with parameters optimized for computational modeling to calculate electric field distributions (simnibs.org) ^50 51^. Seed-based resting-state functional connectivity within and between large-scale task-relevant networks (e.g., frontoparietal and default mode network) will be performed ^39 52 53^ using the CONN toolbox (www.nitrc.org/projects/conn) ^54^. White-matter pathways will be reconstructed from diffusion-weighted images using TRACULA pipeline ^55^ in Freesurfer (http://surfer.nmr.mgh.harvard.edu/), in order to extract tract fractional anisotropy and mean diffusivity ^56-59^. Changes in FA and MD on whole-brain level will be analyzed using FSL’s tract-based spatial statistics (TBSS, www.fmrib.ox.ac.uk/fsl) ^60 61^. MD in gray matter will also be explored for examination of intervention-induced microstructural gray matter change ^62^. Quantifications of metabolite concentrations (such as N-acetylaspartate, glutamate, and gamma-aminobutyric acid, among others) in the MRS voxel will be extracted using software packages in Matlab (The Mathworks Inc., Natick; MA). Segmentation on high-resolution T1 scans will be performed to assess the volume of cortical and subcortical gray matter ^25 63^ using the computational anatomy toolbox (CAT12, http://www.neuro.unijena.de/cat/) and Freesurfer (http://surfer.nmr.mgh.harvard.edu/).

### Retention and Adherence

To ensure adherence and retention throughout the entire study period, participants will be contacted regularly via telephone or email and will be provided with information about their appointments. A few days prior to the intervention, a member of the study team will call the participant to remind them of the upcoming appointments and to check if there are any open questions to address. Additionally, the participants will be provided with a document listing all their appointments with notes regarding parking and caffeine consumption at pre-assessment. At the end of each visit, the participants will be reminded of the date and time of the next session. Throughout the whole study period participants will be encouraged to contact the 24/7 study answer machine if questions or concerns arise. Similarly, they will be prompted to leave a voice message if they are unable to attend or wish to reschedule a session. They then will be contacted to discuss alternative scheduling or to address possible questions and concerns. At the end of the study, participants will receive a reasonable financial reimbursement (approximately 10 € per hour) and the results of their neuropsychological screening and, if they underwent MRI scanning, their structural MRI images on a compact disc. If complete adherence to the protocol is not possible, any effort to collect as much data as possible will be made.

### Data Management and Monitoring

All participant data will be pseudonymized. Spreadsheets containing both participant IDs and personal data will be protected with a password only known to the study staff and will be stored on a secure file server. Similarly, digital data, i.e. output files from computer-based tasks, will be saved on a secure file server directly after acquisition. Non-digitally acquired data will be manually digitalized by a staff member of the research team and double-checked by another member. Progress of data entry and checking procedures will be documented. Files containing subject records will be sorted by participant ID and stored securely. Sensitive data, such as names and medical records, will be scored separately in lockable cabinets in rooms with access restricted to the researchers. Protocols of the tDCS setup of each participant and session will also be stored on the file server and MRI data will be pseudonymized before analysis. Following good scientific practice, data will be stored for at least 10 years.

### Adverse Events Monitoring

Safety and tolerability will be assessed through monitoring any potentially occurring adverse events (AEs) via an adverse events questionnaire ^23^, administered at the end of each training week (V4, V7 and V10). We will refrain from administering the AE questionnaire at every visit, since this would possibly draw the participant’s attention to minor sensations during the stimulation and would only serve as a distraction from the training task. Generally, adverse events during tDCS are rare and minimal. Known AEs with the applied stimulation parameters (20min, 2mA) are skin reddening, a tingling sensation under the electrode sites and occasionally a mild headache ^23^. As we will be using a local anesthetic under the electrode sites, we expect even less noticeable sensations ^64 65^. Investigators will be instructed to monitor AEs and serious AEs (SAEs) throughout the study and document all detected AEs and SAEs. Participants will be informed at baseline assessment about all possible AEs and risks and can withdraw their consent at any time without providing reason. In case a SAE occurs, the study physician will first make an assessment as to whether a causal relationship with the intervention is considered plausible. If more than three of the enrolled participants suffer from SAEs likely to be associated with the intervention (as assessed by the study physician), the trial will be discontinued.

### Statistical Methods

The primary outcome, percent of correct responses in the n-back task at post-compared to the pre-training assessment will be analyzed using ANCOVA with post-assessment n-back performance as dependent variable, pre-assessment n-back performance as covariate, and group allocation (target intervention (n=26) vs. control intervention (n=26)) as independent variable. Linear mixed models will be conducted for secondary outcomes with experimental group as between-subjects factor. All models will be corrected for age and performance at pre-assessment. We will use random intercept models that account for the clustering of measures within individuals. In case of violation of requirements for parametric methods, data will be transformed before analysis. Analyses of primary and secondary outcomes will be reported in detail in the statistical analysis plan to be written and registered before unblinding of investigators performing the analyses. Confirmatory analysis of treatment effects will be conducted within an intention to treat (ITT) framework with multiple imputed data sets in case of missing data (under the assumption of missing completely at random or missing at random). Further as sensitivity analyses, we will perform “per protocol” analyses, including only those participants who finished post-assessment. Feasibility data (% drop-out rate and number of attended training sessions per participant) will be analyzed using descriptive statistics. Data distributions of the feasibility questionnaire items will be visually assessed for normality using q-q plots, and statistically using the Shapiro-Wilk test ^66 67^. Data analysis will be conducted using IBM SPSS Statistics for Windows (IBM Corp., Armonk, NY), MatLab (The Mathworks Inc., 2016), and R software (https://www.R-project.org).

### Patient and public involvement

We will conduct a brief semi-structured interview at the last visit (V12) to assess the patients’ satisfaction with the trial and answer any upcoming questions. The patients were not involved in designing the intervention. All patients will be informed about the study details (e.g. the experimental group they participated in) upon completion of the study.

## Ethics and Dissemination

The study was approved by the ethics committee of the University Medicine of Greifswald and will be conducted in accordance with the Helsinki Declaration. All data collected will be pseudonymized. Results of the study will be made accessible to scientific researchers and health care professionals via publications in peer reviewed journals and presentations at national and international conferences. Furthermore, the scientific and lay public can access the study results on the ClinicalTrails.gov website (Identifier: NCT04817566).

## Conclusion

With this trial, we will for the first time investigate the immediate and long-term effects as well as the feasibility, safety and tolerability of a combined training and brain stimulation intervention on trained (working memory) performance and transfer to other domains as well as on health-related quality of life in patients with post-chemotherapy cognitive impairment. The results of this trial will contribute to the development of non-invasive interventions for a condition affecting everyday lives of many breast cancer patients.

## Data Availability

All data produced in the present study are available upon reasonable request to the authors.

## Protocol amendments

Any substantial amendment to the study protocol will be submitted to the Institutional Ethics Committee for review and approval.

## Consent or assent

A member of the investigational team (study coordinator or study assessor) will collect written informed consent during study enrollment after having reviewed the participant information sheet, participant’s questions, and study inclusion and exclusion criteria.

## Confidentiality

The collected data will be treated as confidential. Direct access to personal information and source data documentation will only be given to study monitors, study assessors, and the research team.

## Availability of data and materials

Anonymized data will be made available to the scientific community upon request.

## Ethics approval and consent to participate

This study was approved by the ethics committee of the University Medicine Greifswald, Germany (BB236/20), date of first approval: 5 January 2021). All procedures conducted during the Neuromod-PCCI trial will be carried out in compliance with the Declaration of Helsinki.

## Authors’ Contributions

AF, DA, and ELB conceptualized and designed this trial. AF and DA are supervising its implementation. AEF and DA are implementing the trial and supervising its conduct. MR, NJ and EK are performing recruitment and assessments. MR, EK, and FT drafted the study protocol. UG, DA and AEF will be performing statistical analyses. All authors will be contributing to the interpretation of the data. All authors read and revised the original draft and consecutive versions of the manuscript. All authors read and approved the final version of the study protocol.

